# Excimer Laser Ablation combined with Drug-coated Balloon for De novo Atherosclerotic Lesions in Lower Extremities

**DOI:** 10.1101/2025.08.01.25332705

**Authors:** Xiaolang Jiang, Lu Yu, Guanyu Qiao, Shuai Ju, Longhua Fan, Hao Liu, Lingwei Zou, Yun Shi, Bin Chen, Junhao Jiang, Tao Ma, Changpo Lin, Gang Fang, Wenqiang Li, Xiaoyan Li, Jianjun Liu, Xin Xu, Daqiao Guo, Weiguo Fu, Zhihui Dong

**Affiliations:** Department of Vascular Surgery, Zhongshan Hospital, Fudan University, Shanghai, 180 Fenglin Road, Shanghai 200032, China; Institute of Vascular Surgery, Fudan University, Shanghai, 180 Fenglin Road, Shanghai 200032, China; National Clinical Research Center for Interventional Medicine, Shanghai, 180 Fenglin Road, Shanghai 200032, China; Department of Vascular and Wound Treatment Center, Jinshan Hospital, Fudan University Shanghai 200540, China; Department of Vascular Surgery, Qingpu Branch of Zhongshan Hospital, Fudan University, 1158 East Gongyuan Road, Shanghai 201715, China

**Keywords:** peripheral artery disease, atherosclerotic obliterans, laser atherectomy, primary patency, technical success

## Abstract

**Background:** The efficacy of excimer laser ablation (ELA) in de novo atherosclerotic lesions of lower extremity artery disease (LEAD) is unknown.

**Objectives:** This real-world study aimed to evaluate the safety and efficacy of ELA combined with drug-coated balloon (DCB) versus DCB alone in LEAD patients.

**Methods:** In this prospective, multicenter, real-world trial (ChiCTR2100051263), patients with de novo atherosclerotic lesions of LEAD were enrolled and allocated to either ELA + DCB or DCB-alone group in a 1:1 ratio. The primary endpoint was 12-month primary patency, with secondary endpoints including technical success, clinically driven target lesion reintervention (CD-TLR), and changes in ankle-brachial index (ABI).

**Results:** A total of 136 patients were enrolled in the study. At baseline, patients in the ELA + DCB group presented significantly higher Rutherford classification (3.7 ± 0.9 *vs*. 4.2 ± 1.0, *p* = 0.007) and longer mean lesion lengths (7.4 ± 2.5 cm *vs.* 8.4 ± 1.9 cm, *p* = 0.012). The ELA + DCB group demonstrated significantly superior 12-month primary patency (87.5% *vs*. 71.2%, *p* = 0.03) and technical success rates (92.7% *vs*. 79.4%, *p* = 0.046) compared to the DCB-alone group. Kaplan-Meier analysis further confirmed sustained patency benefit with ELA + DCB (*p* = 0.015).

**Conclusion:** In this real-world trial, ELA appears to be a promising therapy for LEAD in terms of safety and efficacy. However, these findings need to be corroborated by larger, randomized studies.

**Clinical Trial Registration:** URL: https://www.chictr.org.cn/. Unique identifier: ChiCTR2100051263

## Introduction

The incidence of peripheral arterial disease (PAD) has been increased recently, which is estimated to be >20% in people aged ≥60 years in China ^1,2^. The all-cause mortality in patients with PAD is about 22% ^3^. As the most commonly used methods for treating PAD, percutaneous transluminal angioplasty (PTA) and stenting have been widely used in clinical practice. Despite notable improvements in stenting devices and techniques in the past decade, up to one-half of all patients who receive femoropopliteal stents require secondary interventions (>115,000 procedures annually) due to in-stent restenosis (ISR), a form of arterial recurrent obstruction particularly common in long and complex lesions ^4,5^. The superiority of drug-coated balloon (DCB) in de novo atherosclerotic lesions and ISR has also been demonstrated in previous studies ^6–9^. However, flow-limiting dissection and recoil are still observed in up to 20% of de novo lesions and 16% of ISR cases after the application of DCB, which need to be treated by stenting ^6,9,10^. No matter PTA or stenting, the contents within the vessels are not removed and just squeezing on the vessel wall and have a high risk of recoil and hyperplasia. Although the great saphenous vein bypass surgery demonstrated encouraging long-term outcomes in the BEST-CLI clinical trials, patients who lacked an adequate saphenous vein conduit did not show advantage over endovascular therapy ^11^.

Excimer laser ablation (ELA) could minimize the dissection compared to PTA without debulking, owing to removal of plaque and calcification. Its efficacy in ISR lesions has been demonstrated and also been approved by U.S. Food and Drug Administration (FDA) ^12^. However, its efficacy in de novo atherosclerotic lesions has not been clearly defined. The application of ELA was available in 2019 in our center, and the safety and efficacy of ELA has been preliminarily confirmed in previous studies ^13,14^. Herein, the ELABORATE trial was designed to compare the safety and efficacy of ELA combined with DCB in patients with de novo atherosclerotic obliterans (ASO) in a real-world setting.

## Methods

### Study design

This trial is a prospective, multicenter, real-world trial, which has been registered as a Chinese Clinical Trial (ChiCTR2100051263). Methodology and overall design of the ELABORATE trial have been described previously ^15^; the rationale of ELABORATE was to compare ELA + DCB treatment with DCB alone for improved outcomes. The design of this trial is accordance with the Standard Protocol Items.

The study protocol was approved by the ethics committee at the coordinating center and by the local institutional review board at each participating center. Local primary investigators at each site are responsible for study administration, coordination, and monitoring of this trial at their own centers. An independent data and safety monitoring board oversaw conduct, safety, and efficacy of the trial in scheduled adjudication meetings; data management and statistical analyses were performed by the coordinating center with oversight by members of the ELABORATE executive committee (see Appendix).

### Study population

In brief, consecutive patients at 3 centers in China who had de novo ASO of lower extremities were considered candidates for assignment to ELA + DCB or to DCB alone between September 1, 2021 and the June 30 of 2023. According to the real-world situation, the group participants will be allocated depends on the patients’ request. The double-blind method is also impossible in this trial. The inclusion criteria were age ≥18 years; target lesion length ≤10 cm of the femoropopliteal and below-the-knee (BTK) arteries; significant stenosis ≥70% or chronic total occlusion (CTO) lesions assessed by the angiography; Rutherford class 2–5 with an ankle-brachial index (ABI) <0.9 in the target limb. Patients will be excluded if they meet the following: ISR lesions; acute limb ischemia; simultaneously usage of other debulking devices; serum creatinine >250 umol/L unless dialysis-dependent, contrast agent allergy, coagulation dysfunction, and other severe comorbidities that is impossible to tolerate the procedure.

After 150 patients were evaluated, 136 were enrolled and 130 completed the 12-month follow-up. Written informed consent was obtained from each eligible participant.

### Interventional Procedures

The interventional procedures have been detailed in a previous study ^13,14^. Briefly, for patients in the treatment group, the laser guide (Spectranetics Corp., Colorado Springs, Colorado) will be used to achieve maximum debulking with starting parameter: 45 mJ/mm^2^, 45 Hz, followed by 50–60 mJ/mm^2^ and 50–70 Hz after passage of the lesion.

The necessity of the usage of the distal protective device is at the surgeon’s discretion. For a runoff vessel presenting with (1) long-segment chronic total occlusion (CTO), (2) severely calcified lesions, (3) subacute thrombosis, the use of a distal protective device is recommended during intervention. After ELA in the treatment group, the following procedure is similar in two groups. A standard balloon is used before the DCB. A DCB is selected with 1:1 ratio to the reference vessel diameter (RVD) for 2 min. Angiography is performed at least two different angles to evaluate the minimal luminal diameter (MLD) and dissection (if existed). If a flow-limiting dissection compromising >40% of the lumen or residual stenosis >30% was observed, the bailout stent is used. The distal protective device would be retrieved and the debris >2 mm will be recorded significant ones ^16^. The usage of other debulking devices or any special balloon, such as Chocolate balloon, cutting balloon or Shockwave balloon is prohibited.

The extend of the calcification of the targeted lesions will be assessed according to Peripheral Arterial Calcium Scoring System (PACSS) ^17^. The length, run-off vessels situations, MLD, and the RVD will be visually assessed under the digital subtraction angiography.

### Follow-up strategy

These patients will be given aspirin (100 mg/d) and clopidogrel (75 mg/d) for at least 6 months after discharge and be followed at 3, 6, 12 months and yearly afterwards. The ABI, Doppler ultrasound of the targeted lesions, the ulcers (if existed) will be evaluated at each follow-up point. All participants will be followed for at least 5 years.

### Clinical Outcomes and Endpoints

The primary efficacy outcome is primary patency at 12-month, which is defined as no clinically driven target lesion reintervention (CD-TLR) and peak systolic velocity ratio (PSVR) ≤2.5 at the target lesion assessed by duplex ultrasound. The secondary efficacy outcomes are: (1) CD-TLR; (2) major amputation, defined as amputation above the ankle; (3) technical success, defined as residual stenosis ≤30% and flow-limiting dissection ≤40%; (4) ABI. All the endpoints will be evaluated and recorded by special physicians who are blind to the allocation.

Safety evaluation includes major adverse event (MAE), all-cause mortality through 30-day follow-up, unplanned major amputation, bailout stent, and distal embolization.

### Statistical Analysis

A minimum sample size of 136 patients is determined to be required for this study, assuming 1-sided α = 0.025, β = 80%, assumed 76.1% and 84.9% primary patency at 12 months in the DCB and ELA + DCB groups, respectively, and 25% attrition due to the nature of the real-world study ^12,18^.

Data will be analyzed by SPSS 20.0 (IBM Co., Armonk, NY, USA) and R software (Version 4.3.3). The distribution of continuous variables will be checked firstly. Continuous variables were presented as mean ± standard deviation (SD) with range values for normally distributed data, or as the medians with the interquartile ranges (IQRs) for non-normally distributed data, respectively. Categorical variables were presented as frequencies and percentages. The differences between continuous variables are identified by using 2-sided Student *t* tests or the Wilcoxon rank test. Whereas chi-square test or Fisher exact test is applied to compare the difference between categorical variables. Primary patency and CD-TLR were displayed in a Kaplan-Meier analysis and compared by log-rank analysis. All *p* values are 2-tailed, and *p* <0.05 is defined as statistically significant.

## Results

### Patient enrollment

From September 2021 to June 2023, a total of 150 patients were assessed for eligibility and 136 were enrolled in this prospective cohort. Of the 14 excluded patients, 6 had acute limb ischemia and 8 declined participation. Participants were equally allocated to the DCB-alone group (n = 68) and the ELA + DCB group (n = 68). The final cohort consisted of 103 males (75.7%) with a mean age of 70.9 years (range: 47-93). During the 12-month follow-up, six patients died prior to reaching study endpoints, and they were excluded from the one-year patency and free from CD-TLR analysis. Among these 6 patients, two were in the DCB-alone group, one died due to heart failure and one due to pneumonia. Four patients were in the ELA + DCB group, and they died from pneumonia and myocardial infarction. All remaining participants (n = 130) completed the 12-month follow-up, with no loss to follow-up (**Figure 1**).

**Figure 1.**
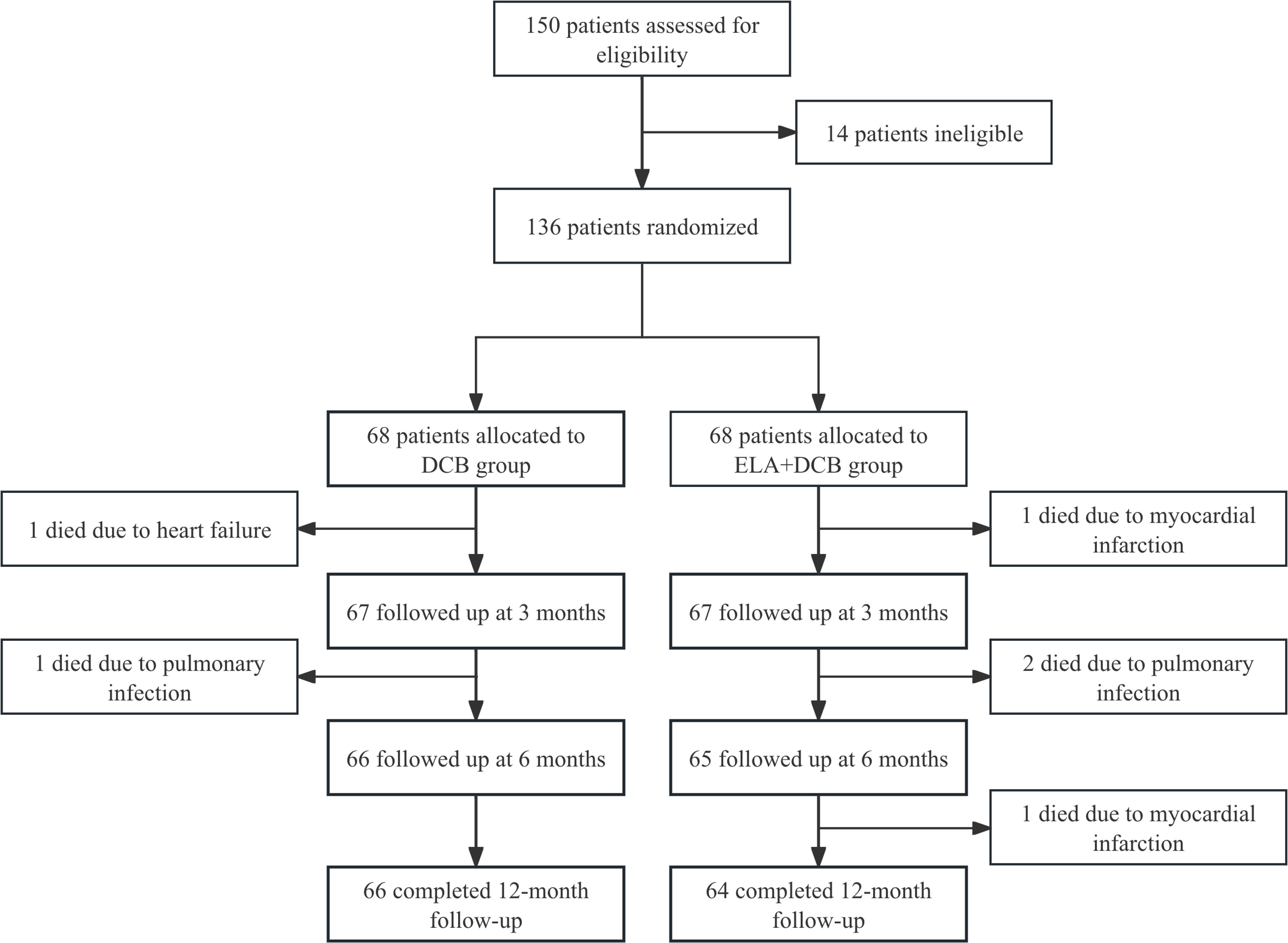
Patient enrollment flowchart. DCB, drug-coated balloon; ELA, excimer laser ablation.

### Baseline characteristics and procedural details

The mean age of patients in the DCB-alone group was 70.5 ± 9.4 years, compared to 71.4 ± 8.5 years in the ELA + DCB group (*p* = 0.442). Males constituted 77.9% (53/68) and 73.5% (50/68) of the DCB-alone and ELA + DCB groups, respectively (*p* = 0.559). Baseline comorbidities and locations of lesions were comparable between the two groups. Preoperative assessments revealed significantly higher Rutherford classification in the ELA+DCB group (4.2 ± 1.0 vs. 3.7 ± 0.9, *p* = 0.007) and longer lesion lengths (8.4 ± 1.9 cm vs. 7.4 ± 2.5 cm, *p* = 0.012). All patients were treated with Orchid paclitaxel-coated peripheral balloon catheter (Acotec Scientific, Beijing, China). The mean length of DCBs used in the two groups were 96.2 ± 24.3 cm and 105.9 ± 18.6 cm, respectively (*p* = 0.010). While the mean diameters of DCBs were comparable (4.3 ± 0.9 cm vs. 4.2 ± 1.0 cm, *p* = 0.557) (**Table 1**).

**Table 1.**
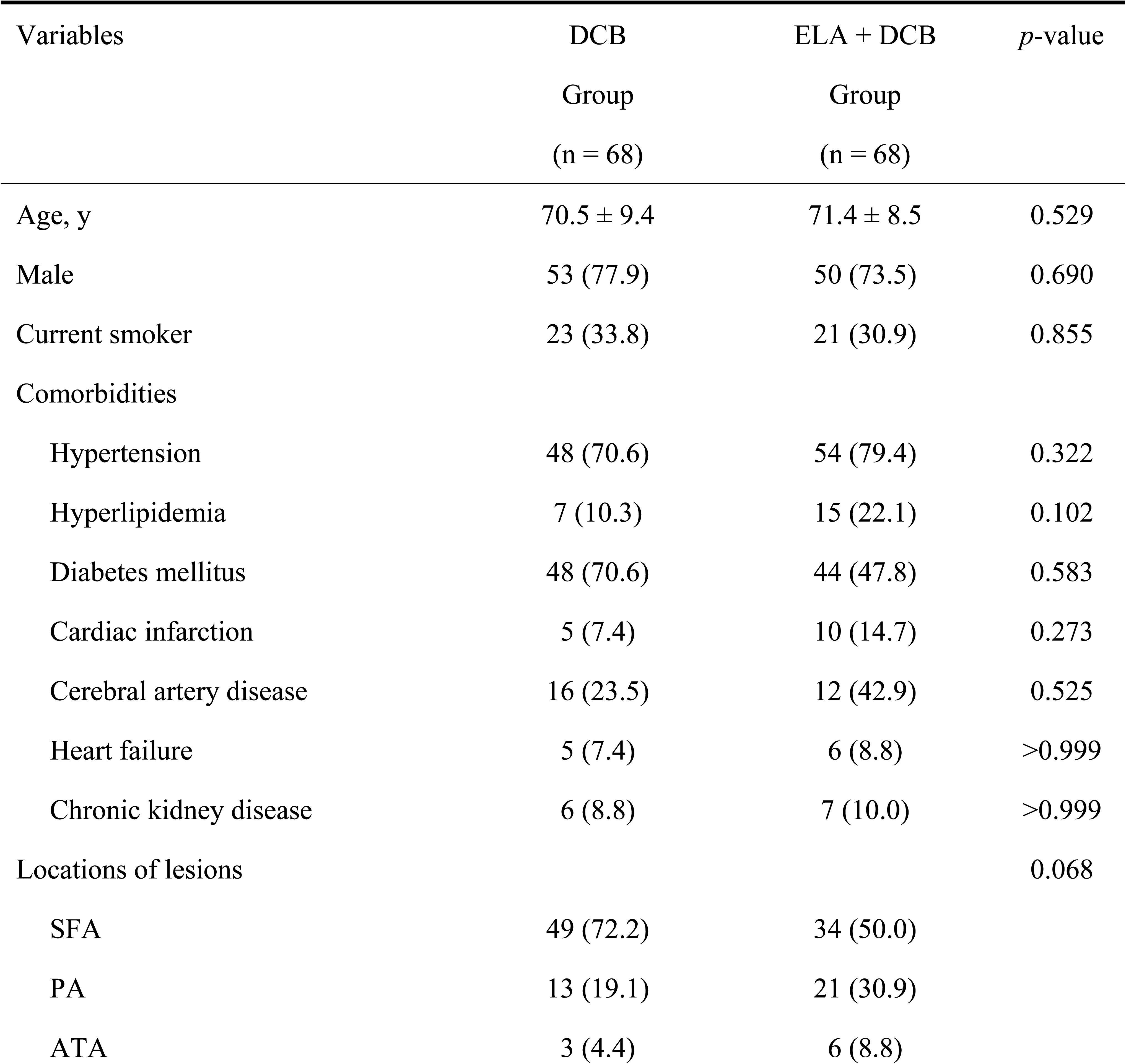
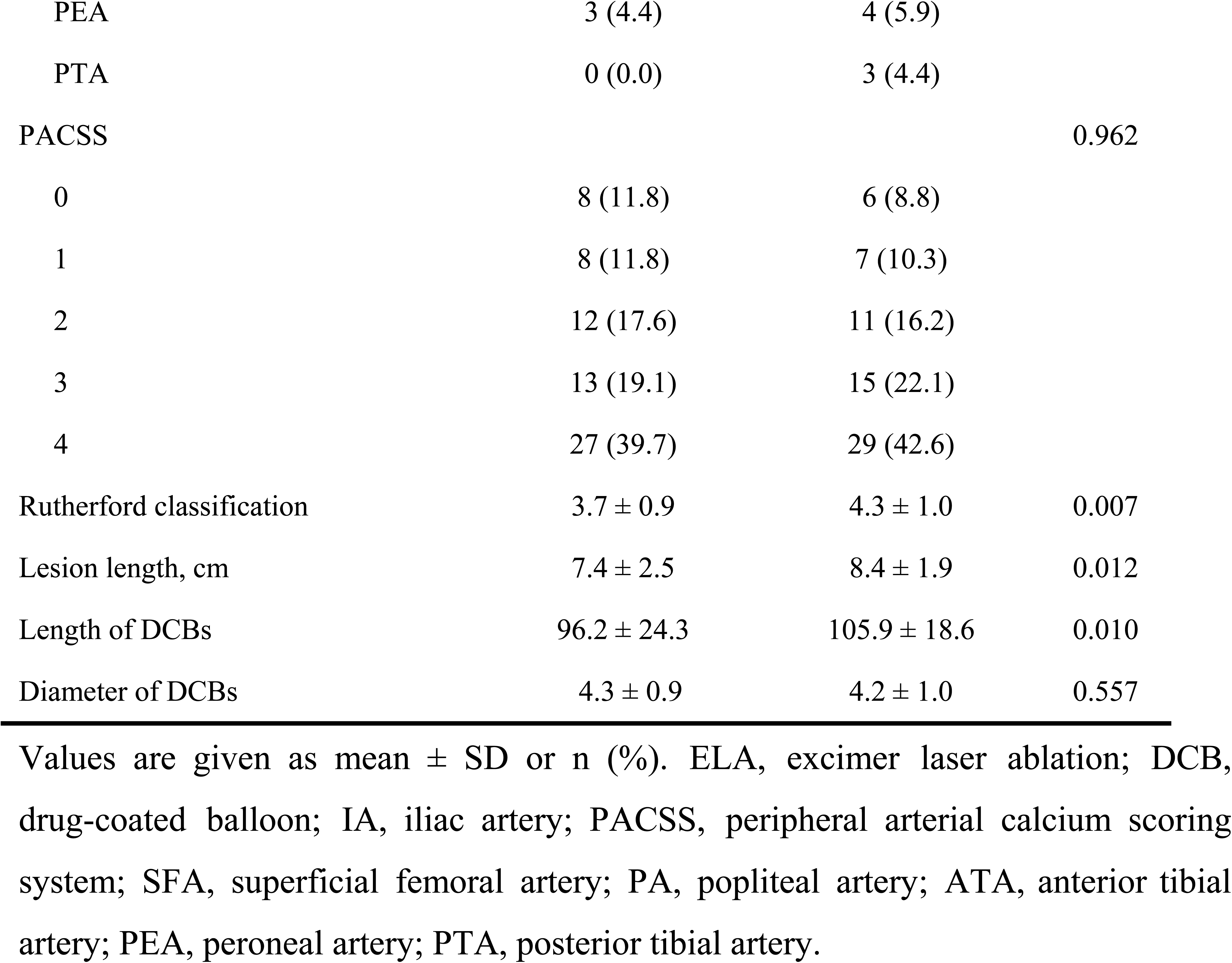
Demographics, lesion characteristics and procedural details.

| Variables | DCB<br>Group<br>(n = 68) | ELA + DCB<br>Group<br>(n = 68) | *p*-value |
| --- | --- | --- | --- |
| Age, y | $70.5 \pm 9.4$ | $71.4 \pm 8.5$ | 0.529 |
| Male | 53 (77.9) | 50 (73.5) | 0.690 |
| Current smoker | 23 (33.8) | 21 (30.9) | 0.855 |
| Comorbidities |  |  |  |
| Hypertension | 48 (70.6) | 54 (79.4) | 0.322 |
| Hyperlipidemia | 7 (10.3) | 15 (22.1) | 0.102 |
| Diabetes mellitus | 48 (70.6) | 44 (47.8) | 0.583 |
| Cardiac infarction | 5 (7.4) | 10 (14.7) | 0.273 |
| Cerebral artery disease | 16 (23.5) | 12 (42.9) | 0.525 |
| Heart failure | 5 (7.4) | 6 (8.8) | >0.999 |
| Chronic kidney disease | 6 (8.8) | 7 (10.0) | >0.999 |
| Locations of lesions |  |  | 0.068 |
| SFA | 49 (72.2) | 34 (50.0) |  |
| PA | 13 (19.1) | 21 (30.9) |  |
| ATA | 3 (4.4) | 6 (8.8) |  |
| PEA | 3 (4.4) | 4 (5.9) |  |
| PTA | 0 (0.0) | 3 (4.4) |  |
| PACSS |  |  | 0.962 |
| 0 | 8 (11.8) | 6 (8.8) |  |
| 1 | 8 (11.8) | 7 (10.3) |  |
| 2 | 12 (17.6) | 11 (16.2) |  |
| 3 | 13 (19.1) | 15 (22.1) |  |
| 4 | 27 (39.7) | 29 (42.6) |  |
| Rutherford classification | 3.7 ± 0.9 | 4.3 ± 1.0 | 0.007 |
| Lesion length, cm | 7.4 ± 2.5 | 8.4 ± 1.9 | 0.012 |
| Length of DCBs | 96.2 ± 24.3 | 105.9 ± 18.6 | 0.010 |
| Diameter of DCBs | 4.3 ± 0.9 | 4.2 ± 1.0 | 0.557 |

Values are given as mean ± SD or n (%). ELA, excimer laser ablation; DCB, drug-coated balloon; IA, iliac artery; PACSS, peripheral arterial calcium scoring system; SFA, superficial femoral artery; PA, popliteal artery; ATA, anterior tibial artery; PEA, peroneal artery; PTA, posterior tibial artery.

### Efficacy outcomes

The 12-month primary patency rate was significantly higher in the ELA + DCB group (87.5%) compared to the DCB-alone group (71.2%, *p* = 0.03) (**Figure 2**). While CD-TLR rates showed a numerical advantage in the ELA + DCB group (10.9% *vs*. 16.7%), but did not reach statistical significance (*p* = 0.448). Notably, primary patency rates at the popliteal segment differed significantly between groups (46.2% vs. 85.0%, *p* = 0.035) (**Table S1**).

**Figure 2.**
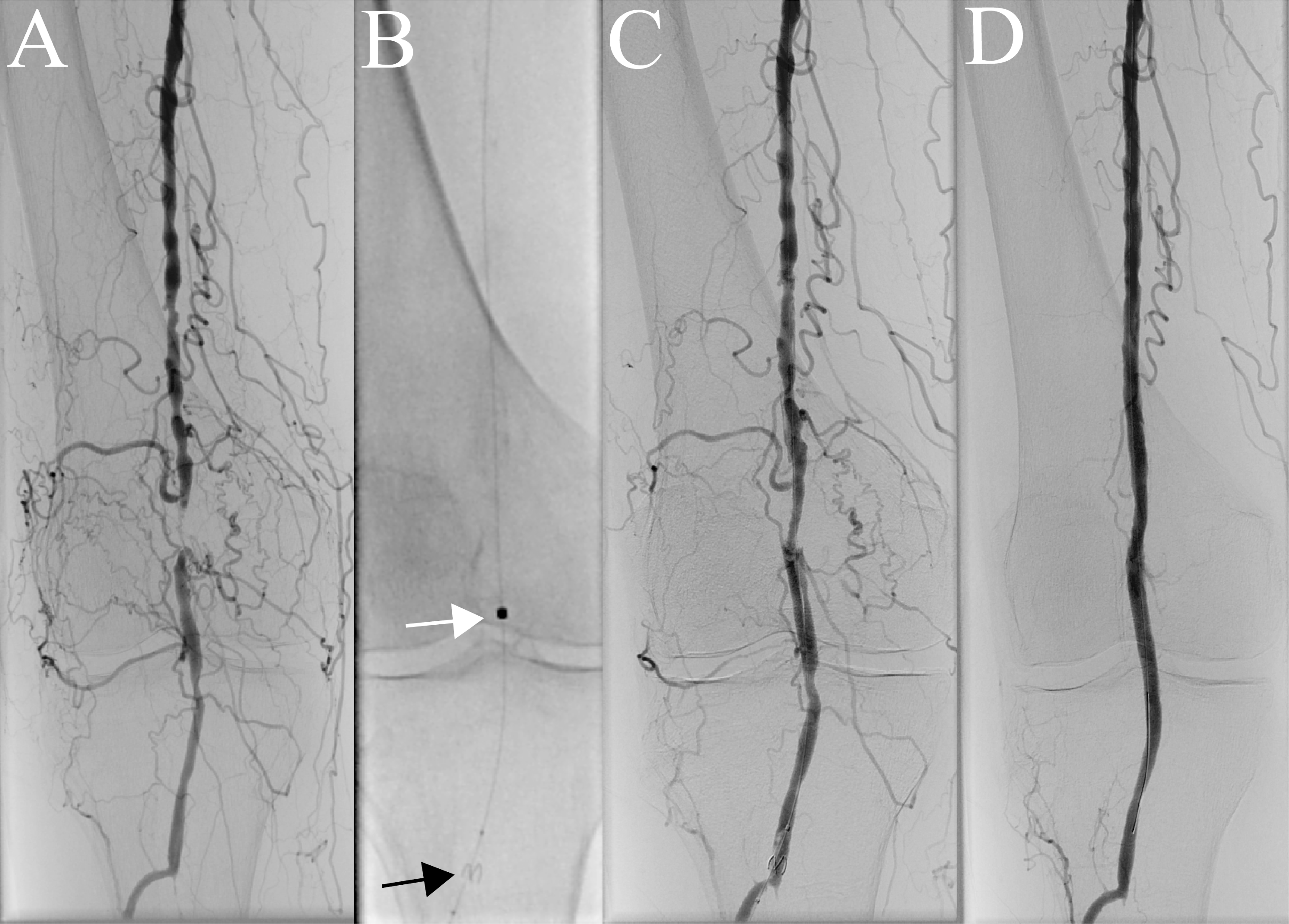
The illustration of treatment of lesions in popliteal artery. (A) The perioperative angiography showed occlusion of the popliteal artery. (B) The distal protective device (black arrow) was applied due to only one runoff to the foot with severe calcification before ELA (white arrow). The ELA was applied for 2 times with an energy of 45-60 mJ and fluence of 60 Hz. (C) The angiography showed satisfactory lumen gain after ELA. (D) Then we applied drug-coated balloons subsequently in the lesion and the completion angiography showed satisfied blood flow and lumen acquirement. DCB, drug-coated balloon; ELA, excimer laser ablation.

Kaplan-Meier analysis demonstrated a more pronounced difference in cumulative primary patency rates between the two groups (*p* = 0.015) (**Figure 3**), whereas freedom from TLR rates remained comparable (*p* = 0.32) (**Figure 4**).

**Figure 3.**
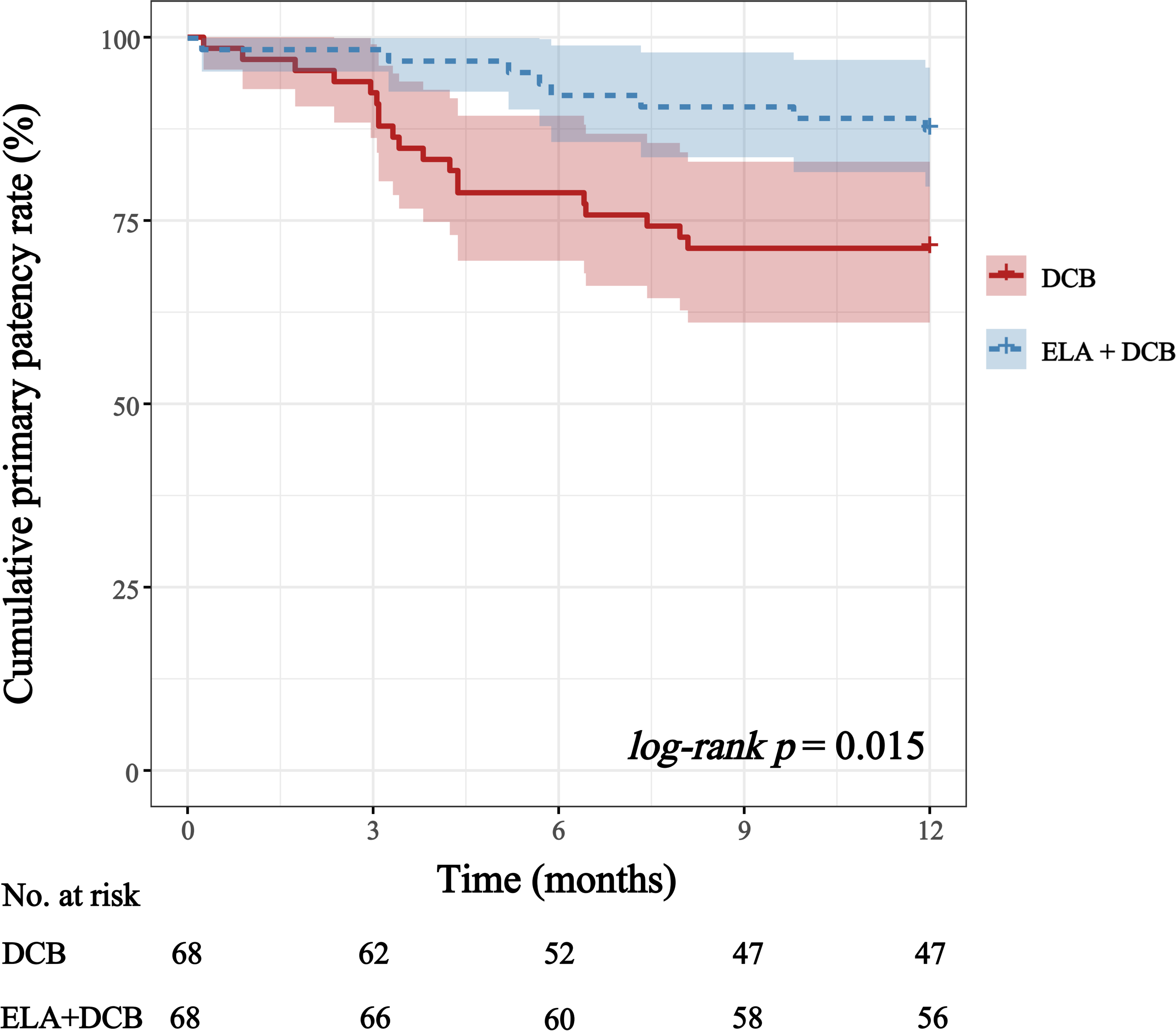
Kaplan-Meier analysis for cumulative primary patency rate in DCB-alone group and ELA + DCB group. The 12-month patency rate was 87.5% in the ELA + DCB group compared with 71.2% in the DCB-alone group. Kaplan-Meier analysis of cumulative patency rates demonstrated a significant difference between the two groups, with a log-rank *p*-value of 0.015. DCB, drug-coated balloon; ELA, excimer laser ablation.

**Figure 4.**
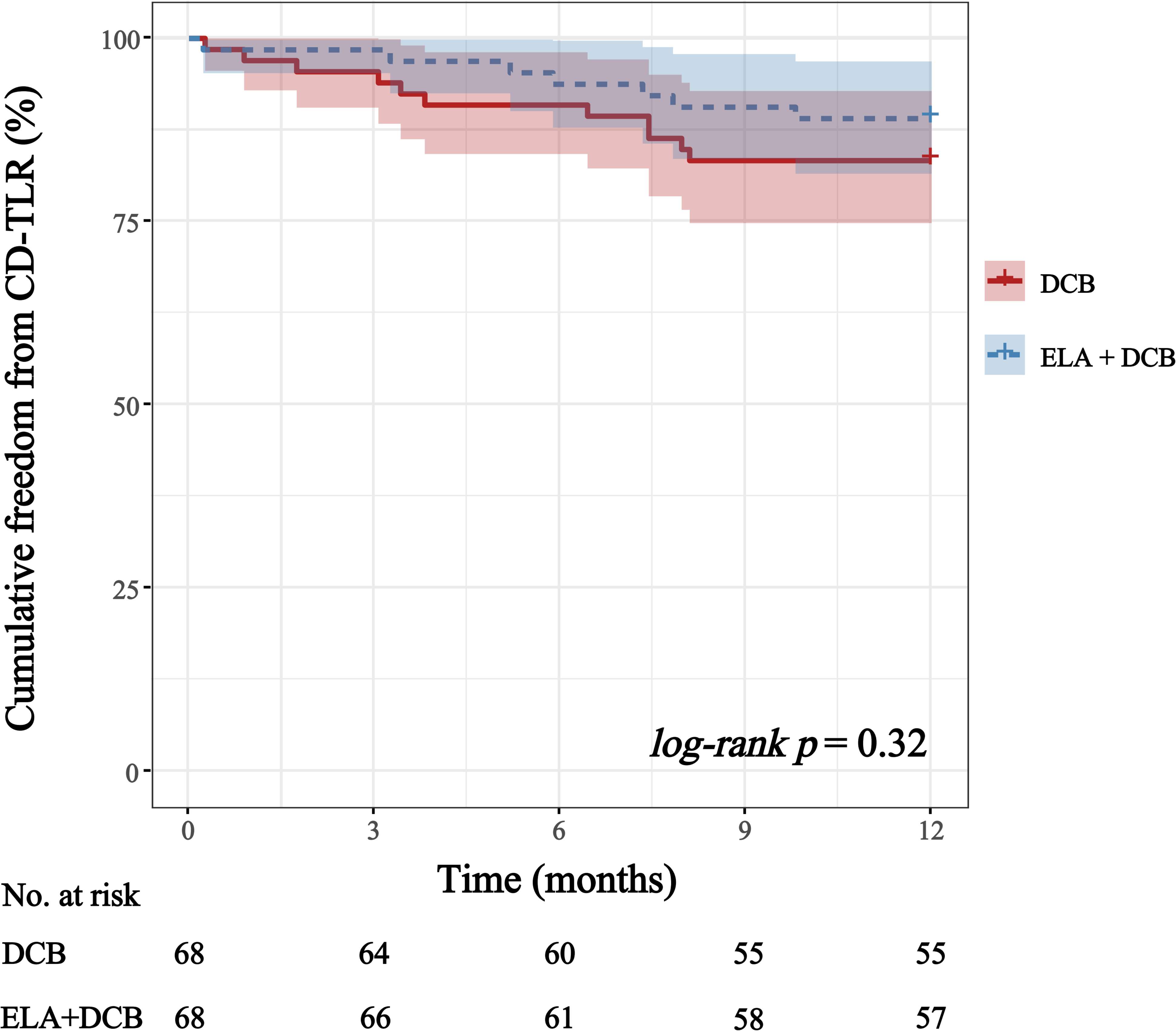
Kaplan-Meier analysis for freedom from CD-TLR rate in DCB-alone group and ELA + DCB group. CD-TLR rate was 10.9% in the ELA + DCB group and 16.7% in the DCB-alone group, with a log-rank *p*-value of 0.32 in Kaplan-Meier analysis. CD-TLR, clinically driven target lesion reintervention; DCB, drug-coated balloon; ELA, excimer laser ablation.

During the 12-month follow-up, unplanned major amputations occurred in 2 patients (2.9%) in the DCB-alone group at 1 and 3 months postoperatively, and 6 patients (8.8%) underwent minor amputations. In the ELA + DCB group, unplanned major amputations were observed in 2 patients (2.9%) at 3 and 11 months postoperatively, and minor amputations in 5 patients (7.4%) (**Table 2**).

**Table 2.** Safety and efficacy outcomes.

|  | DCB<br>Group | ELA + DCB<br>Group | *p*-value |
| --- | --- | --- | --- |
| ***Safety outcomes*** | ***n = 68*** | ***n = 68*** |  |
| Technical success | 54 (79.4) | 63 (92.7) | 0.046 |
| Bailout stent implantation | 13 (19.1) | 5 (7.4) | 0.074 |
| ***Efficacy outcomes*** | ***n = 66*** | ***n = 64*** |  |
| 12-month primary patency | 47 (71.2) | 56 (87.5) | 0.030 |
| CD-TLR | 11 (16.7) | 7 (10.9) | 0.448 |
| Unplanned major amputation | 2 (3.0) | 2 (3.1) | 0.975 |
| Minor amputation | 6 (9.1) | 5 (7.8) | 0.793 |

Values are given as mean ± SD or n (%). DCB, drug-coated balloon; ELA, excimer laser ablation; CD-TLR, clinically driven target lesion reintervention.

**Table 3.** Changes in ABI before and after intervention.

| Time Point | DCB Group<br>(n = 68) | ELA + DCB Group<br>(n = 68) |
| --- | --- | --- |
| Preoperative | $0.52 \pm 0.17$ | $0.50 \pm 0.15$ |
| Immediate postoperative | $0.93 \pm 0.13$ | $0.91 \pm 0.17$ |
| 3 Months | $0.88 \pm 0.22$ | $0.89 \pm 0.19$ |
| 6 Months | $0.83 \pm 0.27$ | $0.81 \pm 0.24$ |
| 12 Months | $0.82 \pm 0.20$ | $0.80 \pm 0.27$ |

Values are given as mean ± SD. ABI, ankle-brachial index; DCB, drug-coated balloon; ELA, excimer laser ablation.

### Safety outcomes

The technical success rate was significantly higher in the ELA + DCB group (92.7%) compared to the DCB-alone group (79.4%, *p* = 0.046). Bailout stent implantation was required in 19.1% (13/68) of the DCB-alone group versus 7.4% (5/68) of the ELA + DCB group (*p* = 0.074).

Among the 14 technical failures in the DCB-alone group, bailout stents were deployed in 13 cases: 11 cases due to flow-limiting dissections and 2 cases attributed to elastic recoil of the vessel. In contrast, all 5 technical failures in the ELA + DCB group were exclusively associated with flow-limiting dissections, necessitating stent implantation.

Distal protective devices were utilized in 10 patients (14.7%) in the ELA + DCB group. No instances of distal embolization occurred during the procedures in either group.

## Discussion

As the first real-world setting prospective clinical study, the ELABORATE trial showed that ELA + DCB achieved superior efficacy and safety compared with DCB alone. Generally, this trial demonstrated that ELA with adjunctive DCB is favorable in the treatment of de novo atherosclerotic lesions in the LEAD, with significant improvements in technical success compared with DCB alone. The primary patency remained significantly better in the ELA + DCB group as compared with DCB alone throughout the study.

ELA may be advantageous for ISR patients because of the ability of this technique to effectively remove hyperplastic tissue ^19^. The predicate device to the Turbo Tandem laser catheter showed significant plaque removal and increased vessel compliance in de novo lesions of the femoropopliteal artery ^20^. DCB has shown superior prevention of restenosis in de novo and ISR lesions of the femoropopliteal artery compared with PTA alone ^18^; however, challenges still existed in severely calcified lesions, which were proven to be independent risk factors of clinical outcomes of DCB ^21^. Because the vessel calcification may act as a physical barrier to optimal drug penetration and adequate distribution after DCB. Excimer lasing re-opens and enlarges the lumen and smoothens the lumen wall, enhancing the angiographic result of adjunctive DCB. Meanwhile, ELA eliminates the calcification and allows the drugs of DCB penetrating into the vessel wall, thus enhancing the clinical effects. The calcifications in two groups were comparable, but the technical success (92.7% vs. 79.4%, *p* = 0.046) and primary patency (87.5% vs. 71.2%, *p* = 0.03) was significantly favorable in ELA + DCB group.

The ELA + DCB group demonstrated significantly higher technical success rates, thereby reducing bailout stenting—an especially valuable outcome for popliteal artery interventions. The consensus that avoidance of deployment of stents in popliteal artery has been reached ^22,23^. Because the popliteal artery is exposed to some of the most brutal forces seen in the lower limb during normal knee flexion ^24^. Therefore, the balloon-based angioplasty, especially DCB, was the main treatment in these lesions ^25^. However, balloon angioplasty alone functions by creating dissections, and postangioplasty dissection is associated with higher technical failure, worse patency, and more reinterventions ^8,26^. Hence, it is crucial for vessel preparation to minimize flow-limiting dissections and to maximize the luminal gain before using DCB. ELA treatment has previously demonstrated efficacy and safety in the treatment of lesions in popliteal artery ^27^. The EXCITE ISR trial demonstrated the superiority of ELA + PTA over PTA alone for treating femoropopliteal ISR lesions ^12^. However, as a RCT with stringent inclusion and exclusion criteria, its patient population was inherently limited. Moreover, its primary efficacy endpoint was TLR at 6-month follow-up, lacking long-term outcome data. Our real-world study enrolled patients better representing the actual clinical disease spectrum, making its findings more generalizable to routine practice. The hypothesis that ELA debulking with adjunctive DCB could improve drug delivery and reduce procedural complications and bailout stenting was confirmed in this study. Among the enrolled patients in this trial, 21 (30.9%) with popliteal lesions received ELA + DCB versus 13 (19.1%) treated with DCB alone, with superior technical success and primary patency in the ELA + DCB group. The higher patency rates and lower bailout stenting requirements observed with ELA + DCB hold significant clinical implications, offering a promising therapeutic alternative for the popliteal artery. However, the current findings require confirmation in larger cohorts due to the limited sample size.

Notably, the ELA + DCB group showed a numerically lower but statistically non-significant reduction in CD-TLR rates compared to DCB-alone (10.9% vs. 16.7%, *p* = 0.448), with Kaplan-Meier analysis similarly demonstrating no significant difference (*p* = 0.32). This outcome may be related to the different management of restenosis between the two groups: among the 19 restenosis cases in the DCB-alone group, only 11 (57.9%) underwent reintervention due to clinical symptoms, whereas 7 of the 8 restenosis cases (87.5%) in the ELA + DCB group received CD-TLR.

While randomized controlled trial (RCT) ensures high internal validity, their strict eligibility criteria limits real-world applicability^12,28^. Retrospective registries capture broader populations but suffer from data heterogeneity and selection bias. Well-designed real-world studies like ELABORATE bridge this gap, balancing scientific rigor with clinical relevance.^29^. By aligning inclusion criteria with real-world practice and limiting exclusions to absolute contraindications, ELABORATE maintains standardization while achieving superior external validity through unselected patient populations and routine practice patterns.

Procedural complications were both low in two groups. ELA + DCB treatment group showed significantly fewer bailout stents. Distal embolization could not be neglect in the usage of debulking device. The usage of directional atherectomy (DA) was proven to be the risk factors of distal embolization and distal protective device was routinely recommended^30,31^. However, the distal protective device was not recommended in the usage of ELA. Because the contents in the lumen is lasered into small debris with a diameter of <25μm, which was smaller than the diameter of white blood cells. Also in the DEEP EMBOLI trial, Shammas et al found that the incidence of debris was 22%, which was equal to that in the stenting group^30^. Despite all these, the incidence of embolization was reported to be 3.6%-22.0%^32,33^.

In our earlier clinical practice, we did not use the distal protection, and the distal embolization occurred in 5.6% of cases ^14^. In this trial, distal protective devices were selectively deployed in 10 high-risk ELA procedures, with six cases involving long-segment CTOs and four cases featuring severely calcified lesions. Significant debris exceeding 2 mm was captured in three cases of severe calcification. No instance of distal embolization was observed in this trial. This marked improvement reflects our progressive refinement in ELA application, including enhanced patient selection based on lesion characteristics, targeted use of embolic protection in anatomically high-risk subgroups, and accumulated technical expertise from prolonged procedural experience.

## Study limitations

The present study had limitations. First, the observational nature of this real-world study precluded blinding of both participants and clinicians, potentially introducing selection bias and confounding factors that cannot be fully eliminated. Second, modest sample size constrained statistical power and subgroup analyses. Third, the current study intentionally restricted lesion length to ≤10 cm. This decision was based on the technical characteristics of ELA, where slower advancement rates (approximately 1 mm/min) raise concerns about prolonged procedure times and increased risks of flow-limiting dissections and distal embolization in longer lesions. This anatomic rationale aligns with the EXCITE ISR Trial, which restricted peri-stent lesions to ≤3 cm in length to minimize thermal injury risks during laser-assisted interventions for in-stent restenosis^12^. While this selection criterion ensured procedural safety during initial evaluation, it may limit generalizability to patients with more extensive disease. In the future, we will enroll patients with lesions up to 20 cm, which will provide critical real-world evidence on the safety and efficacy of ELA + DCB in LEAD.

## Conclusion

This prospective real-world study provides preliminary clinical evidence supporting the efficacy of ELA as an adjunct to DCB angioplasty for de novo atherosclerotic lesions in LEAD. Compared with DCB alone, ELA + DCB demonstrated significantly superior 12-month primary patency and higher procedural success rates, reinforcing its potential as an optimized endovascular strategy for LEAD lesions. These findings warrant validation with extended follow-up periods and well-designed randomized studies.

## Ethics approval and consent to participate

This study design was approved by the Ethics Committee for the Protection of Human Subjects at Zhongshan Hospital, Fudan University, Shanghai, China. All included patients were informed about the nature of the study and gave their written informed consent.

## Consent for publication

All patients signed a consent form for their data to be used for research or publication.

## Abbreviation

ABI: Ankle-Brachial Index
ASO: Atherosclerotic Obliterans
BTK: Below-The-Knee
CD-TLR: Clinically Driven Target Lesion Reintervention
ChiCTR: Chinese Clinical Trial Registry
CTO: Chronic Total Occlusion
DA: Directional Atherectomy
DCB: Drug-Coated Balloon
ELA: Excimer Laser Ablation
FDA U.S.: Food and Drug Administration
ISR: In-Stent Restenosis
LEAD: Lower Extremity Artery Disease MAE Major Adverse Event
MLD: Minimal Luminal Diameter PAD Peripheral Arterial Disease
PACSS: Peripheral Arterial Calcium Scoring System
PSVR: Peak Systolic Velocity Ratio
PTA: Percutaneous Transluminal Angioplasty
RCT: Randomized Controlled Trial
RVD: Reference Vessel Diameter

## Data Availability

The datasets analyzed during this study are available from the corresponding author on reasonable request.

---

**Supplementary Table 1.** Patency rates by lesion location.

|  | DCB Group<br>(n = 66) | ELA + DCB Group<br>(n = 64) | *p*-value |
| --- | --- | --- | --- |
| 12-month primary patency | 71.2% (47/66) | 87.5% (56/64) | 0.030 |
| SFA | 80.9% (38/47) | 90.9% (30/33) | 0.214 |
| PA | 46.2% (6/13) | 85.0% (17/20) | 0.035 |
| BTK | 50.0% (3/6) | 81.2% (9/11) | 0.419 |

DCB, drug-coated balloon; ELA, excimer laser ablation; SFA, superficial femoral artery; PA, popliteal artery; BTK, below-the-knee.

